# Convergent genetic and functional evidence implicate *KCND3* in Brugada Syndrome

**DOI:** 10.64898/2026.09.30.26364181

**Authors:** Henan Liu, Arie O. Verkerk, Pierre Lindenbaum, Isabella Mengarelli, Fenna Tuijnenburg, Rezbieara P. Rahman, John Mauleekoonphairoj, Maarten Kooyman, Dominic Zimmerman, Estelle Baron, Ingrid Krapels, Paul G. Volders, Leander Beekman, Laura Cazón Varela, Amalio Ruiz Salas, Rafik Tadros, Ammar Al-Chalabi, Jan H. Veldink, Aurélie Thollet, Eric Charpentier, Philippe Mabo, Pascale Guicheney, Antoine Leenhardt, Arjan C. Houweling, Hanno L. Tan, Jean François Deleuze, France GEN-REF consortium, Juan Pablo Ochoa, Ruben Coronel, Ronald Wilders, Yanushi D. Wijeyeratne, Wanwarang Wongcharoen, Pakorn Chandanamattha, Pieter G. Postema, Jean-Baptiste Gourraud, Jean-Jacques Schott, Christian Dina, Gumpanart Veerakul, Frederic Sacher, Jacques Mansourati, Ahmad S. Amin, Vincent Probst, Arthur A. Wilde, Richard Redon, Pier D. Lambiase, Elijah R. Behr, Apichai Khongphatthanayothin, Koonlawee Nademanee, Julien Barc, Roddy Walsh, Connie R. Bezzina

**Affiliations:** Department of Experimental Cardiology, Amsterdam University Medical Centre, University of Amsterdam, Amsterdam, The Netherlands; Amsterdam Cardiovascular Sciences, Cardiomyopathy and Arrhythmia, Amsterdam, The Netherlands; European Reference Network for Rare, Low Prevalence and Complex Diseases of the Heart: ERN GUARD Heart; Nantes Université, CHU Nantes, CNRS, INSERM, l’Institut du Thorax, F-44000 Nantes, France; Department of Clinical Cardiology, Heart Centre, Amsterdam University Medical Centre, location AMC, The Netherlands; Cardiovascular Research Institute, City St George’s University of London, London, UK; Centre of Excellence in Arrhythmia Research, Department of Medicine, Faculty of Medicine, Chulalongkorn University, Bangkok, Thailand; Department of Clinical Genetics, Maastricht University Medical Centre+, Maastricht, The Netherlands; Department of Cardiology, CARIM, Maastricht University Medical Centre, Maastricht, The Netherlands; Health in Code S.L., A Coruña, Spain; Department of Medicine, Heart Area, Hospital Universitario Virgen de la Victoria, IBIMA Plataforma BIONAND (Instituto de Investigación Biomédica de Málaga y Plataforma en Nanomedicina), CIBERCV (Centro de Investigación Biomédica en Red Enfermedades Cardiovasculares), Universidad de Málaga (UMA), 29010 Málaga, Spain; Centro de Investigación Biomédica en Red de Enfermedades Cardiovasculares (CIBERCV), 28029 Madrid, Spain; Cardiovascular Genetics Centre, Montreal Heart Institute and Faculty of Medicine, Université de Montréal, Montreal, Canada; Department of Neurology, King’s College London, London, UK; Department of Neurology, UMC Utrecht Brain Centre, University Medical Centre Utrecht, Utrecht University, Utrecht, The Netherlands; Service de Cardiologie, CHU de Rennes, Rennes, France; INSERM, Sorbonne University, UMRS 1166, Institute of Cardiometabolism and Nutrition (ICAN), Paris, France; CNMR Maladies Cardiaques Héréditaires Rares, Hôpital Bichat, Université de Paris, Paris, France; AP-HP, Service de Cardiologie, Hôpital Bichat, Paris, France; Department of Human Genetics, Amsterdam UMC, Amsterdam, The Netherlands; Centre National de Recherche en Génomique Humaine (CNRGH), Institut de Biologie François Jacob, CEA, Université Paris-Saclay, Evry, France; Laboratory of Excellence GENMED (Medical Genomics), Paris, France; Centre d’Etude du Polymorphisme Humain, Fondation Jean Dausset, Paris, France; Spanish National Centre for Cardiovascular Research (CNIC), Madrid, Spain; Department of Medical Biology, Amsterdam Cardiovascular Sciences, Amsterdam University Medical Centre, University of Amsterdam, 1105 AZ Amsterdam, The Netherlands; Cardiovascular Clinical Academic Group, St George’s University Hospitals NHS Foundation Trust, London, UK; Department of Medicine, Faculty of Medicine, Chiang Mai University, Chiang Mai, Thailand; Department of Medicine, Faculty of Medicine at Ramathibodi Hospital, Mahidol University, Bangkok, Thailand; Bangkok Heart Hospital, Bangkok, Thailand; IHU Liryc, Electrophysiology and Heart Modeling Institute, fondation Bordeaux Université, Pessac-Bordeaux, France; Université Bordeaux, Centre de recherche Cardio-Thoracique de Bordeaux, Bordeaux, France; INSERM, Centre de recherche Cardio-Thoracique de Bordeaux, Bordeaux, France; Electrophysiology and Ablation Unit, Bordeaux University Hospital (CHU), Pessac, France; Department of Cardiology, University Hospital of Brest, Brest, France; Institute of Cardiovascular Science, University College London (UCL) and Barts Heart Centre, London, UK; Heart Institute, Bumrungraj Hospital, Bangkok, Thailand; Pacific Rim Electrophysiology Research Institute, Las Vegas, Nevada, USA

**Author notes:** These authors contributed equally to this work (Henan Liu, Arie O. Verkerk). These authors co-supervised this work (Julien Barc, Roddy Walsh, Connie R. Bezzina). **Corresponding author:** Connie R. Bezzina, Department of Experimental Cardiology, Amsterdam University Medical Centre, University of Amsterdam, Room K2-116, Meibergdreef 9, 1105 AZ Amsterdam, The Netherlands.

**Keywords:** Brugada Syndrome, sudden cardiac death, epistasis, *KCND3*, K_v_4.3, transient outward potassium current (*I*_to_)

## Abstract

**Background:** Dysfunction of the *KCND3*-encoded K_v_4.3 channel, which carries the transient outward potassium current (*I*_*to*_), has long been suspected to contribute to the pathophysiology of Brugada Syndrome (BrS). Although recent genome-wide association studies (GWAS) have implicated *KCND3*, the role of rare variation in this gene remains uncertain and clinically unactionable.

**Methods:** Genome sequencing was performed in 983 BrS probands from Europe and Thailand. Putative gain-of-function (GoF) rare variants in *KCND3* identified in patients were functionally characterized by patch-clamp analysis of K_v_4.3 current properties using a HEK293 cell heterozygous model. Biobank datasets were utilized to evaluate rare-common variant interaction at the *KCND3* locus.

**Results:** Eight BrS cases carried six different rare *KCND3* variants which clustered at the K_v_4.3 C-terminus, with terminal exon rare variants significantly enriched in cases versus study (p=0.004) and population (p=2.0e^−6^) controls. Four of the five *KCND3* case variants studied demonstrated significant GoF effects versus wild-type and putative benign variants, including increased *I*_*to*_ amplitude (p.Leu586Val, p.Ala634Thr) and slower inactivation (p.Gly600Arg, p.Ala654Thr, p.Ala634Thr). Analysis of p.Gly600Arg carriers in the UK Biobank and *All of Us* revealed a significant enrichment of the protective *KCND3* expression-reducing allele of the GWAS lead single-nucleotide polymorphism rs12090194, suggesting a possible epistatic common-rare variant interaction modifies penetrance in carriers.

**Conclusions:** GoF rare variants at the C-terminus of *KCND3* are associated with BrS in a small proportion of cases. Determination of the rare and common variant status in BrS patients and their relatives may provide information about disease risk.

## Introduction

Brugada Syndrome (BrS) is a disorder presenting with distinctive J-wave augmentation and coved-type ST elevation in the right precordial electrocardiogram (ECG) leads, alongside elevated risk of malignant ventricular arrhythmias and sudden cardiac death (SCD) ^1,2^. It forms an entity within the J-wave syndrome spectrum of disorders, which also encompasses early repolarization syndrome (ERS) with which it shares electrocardiographic and clinical features ^3–5^. While originally viewed as a purely electrical disorder, BrS is now recognized along a microstructural–electrical continuum, with sodium channel dysfunction and subtle epicardial fibrosis of the right ventricular outflow tract as key contributors ^5,6^.

BrS has a complex genetic architecture, whereby an individual’s disease risk is determined by the cumulative effects of multiple genetic variants across a spectrum of population frequencies and effect sizes. Rare, large-effect coding variants in *SCN5A* that result in loss of function of the encoded Na_V_1.5 channel are identified in approximately 20% of European patients and 5–10% of Asian patients ^7,8^. At the other end of the spectrum, genome-wide association studies (GWAS) conducted in European and Asian populations have identified 27 chromosomal loci harbouring common small-effect susceptibility variants ^9–13^.

Notably, aside from *SCN5A* at the chromosome 3 GWAS locus, *KCND3* at the chromosome 1 locus is the only other ion channel gene prioritized at the BrS-associated GWAS loci. *KCND3* encodes the pore-forming α-subunit of the tetrameric K_v_4.3 channel, carrying the transient outward current (*I*_*to*_) in human cardiomyocytes, which plays a critical role in the early repolarization phase of the ventricular action potential. A role for K_v_4.3 has long been hypothesized for BrS. The repolarization hypothesis attributes the electrocardiographic and arrhythmogenic characteristics of the disorder to regionally accentuated differences in the density of *I*_*to*_ within the right ventricle ^14^. In a second proposed mechanism, increased *I*_*to*_ may contribute to impaired depolarization and conduction slowing - to which the right ventricle is exquisitely sensitive - by opposing inward depolarizing currents ^15,16^.

Rare missense variants in *KCND3* have also been implicated in BrS, with 2 studies describing cases with variants that were demonstrated to have GoF effects (increased *I*_*to*_ density and altered gating kinetics) in heterologous cell assays ^17,18^. However, the ClinGen re-evaluation of gene-disease evidence for BrS in 2018 concluded that *KCND3* (like all implicated genes except for *SCN5A*) had disputed association, citing limited cases/replication, the lack of statistically significant genetic evidence (segregation in large family pedigrees or rare variant case-control enrichment) and discrepancies between *in vitro* functional data and the population frequencies of reported variants ^19^. However, as the ClinGen framework is based on Mendelian models of genetic disease, it may not account for the increasingly complex genetic architecture of BrS.

Here, we used genome sequence data from 983 BrS probands to reassess the potential role of rare/low-frequency variants in *KCND3*, providing genetic and functional evidence supporting a contribution to disease susceptibility in the setting of complex inheritance.

## Methods

For detailed methods, please refer to the supplementary materials.

## Results

### Rare C-terminal KCND3 variants in BrS patients

*KCND3* was sequenced by genome sequencing in 752 BrS probands of European ancestry (from the Netherlands, France and the UK) and 231 BrS probands from Thailand. Genome sequencing data from 1,827 European-ancestry and 500 Thai ancestry individuals was utilized as study controls. All BrS cases were diagnosed according to international guidelines ^20–22^, with ECGs reviewed by BrS-expert electrophysiologists (**Supplemental Methods**). Six rare (gnomAD v4.1 filtering allele frequency < 0.0001) missense variants were detected in eight cases (**Table S1**). While there was no overall rare variant enrichment compared to ancestry-matched study controls (0.81% vs. 0.52%, p=0.33), the case variants were notably clustered at the C-terminus of the protein (**Figure 1A**). Seven of the eight cases harboured variants in the last exon, where a significant 8-fold case-control enrichment was observed (p=0.004 vs. study controls, p=2.0e^−6^ vs. gnomAD population controls) with consistent effects for both European and Thai/East Asian groups (**Figure 1B, Table S2**).

**Figure 1.**
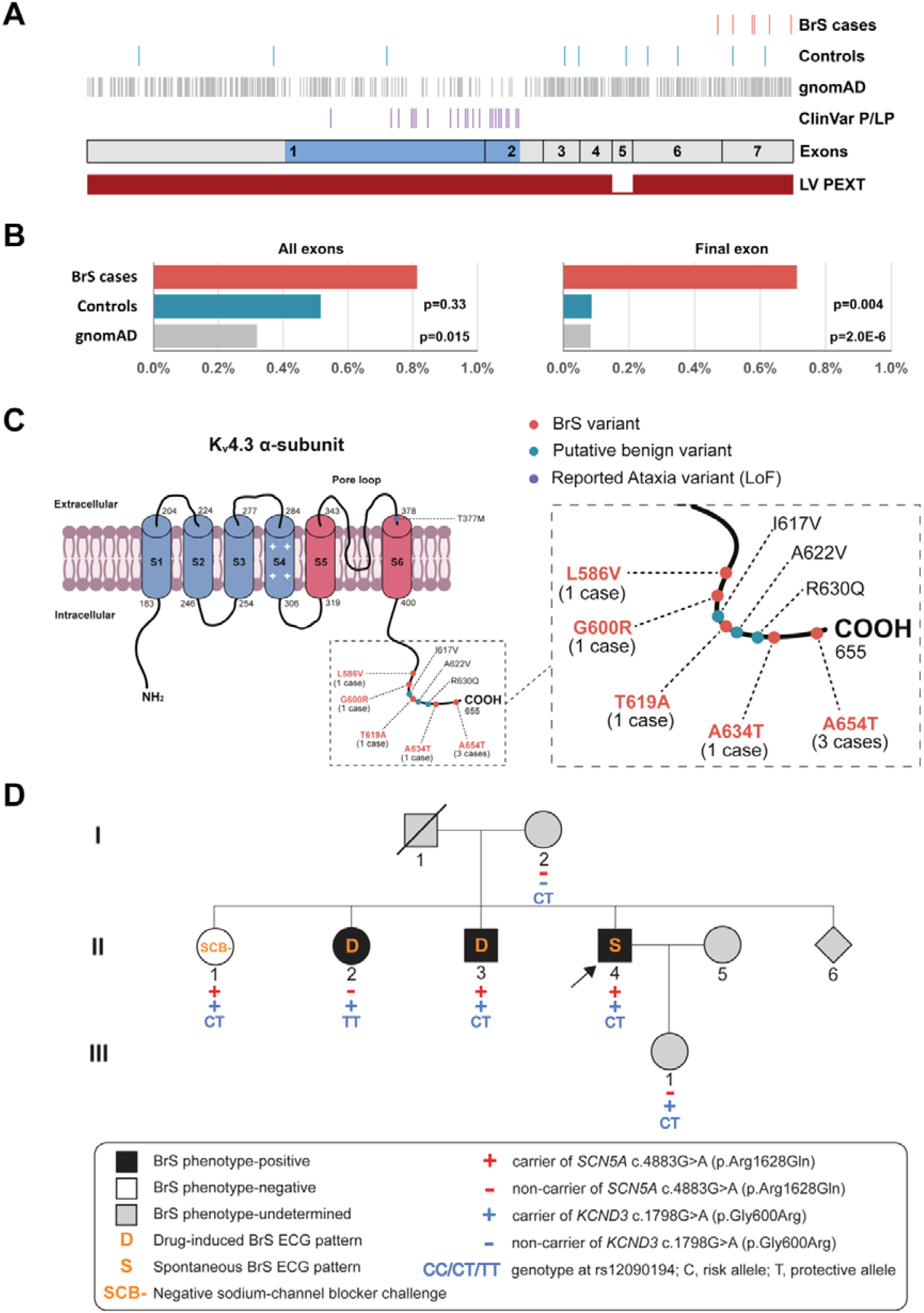
Location of rare missense variants in *KCND3* transcript and K_v_4.3 protein. **(A)**Location of rare (gnomAD v4.1 filtering allele frequency < 0.0001) missense and inframe indel variants with respect to the seven coding exons of the ENST00000302127 / NM_001378969 transcript for: BrS cases in this study, controls in this study, gnomAD v4.1 exomes, pathogenic / likely pathogenic (P/LP) variants in ClinVar (largely associated with spinocerebellar ataxia). The transmembrane region is highlighted in blue. The final track shows the proportion expressed across transcripts (PEXT) score, highlighting which exons are included in transcripts expressed in the left ventricle (LV) (data from gnomAD/GTEx). The canonical transcript including exon 5 appears to be relatively lowly expressed in cardiac tissue. **(B)**Percentage of individuals with rare (gnomAD v4.1 filtering allele frequency < 0.0001) *KCND3* missense variants in BrS (n=983), controls (n=2327) and gnomAD (n=728290), across all coding exons (left) and final exon only (right). **(C)**Schematic diagram of the K_v_4.3 channel pore-forming α-subunit showing positions of the nine *KCND3* missense variants analysed in the present study. The six transmembrane domains (S1–S6), pore loop (between S5 and S6), and intracellular N- and C-termini are illustrated. The dashed box indicates a magnified view of the C-terminal region, where eight of the nine variants are clustered. LoF = loss-of-function. **(D)**Pedigree of the family harbouring *KCND3*:p.Gly600Arg. Squares and circles indicate males and females, respectively; a diagonal slash denotes a deceased individual, and the arrow indicates the proband. Black, white, and grey symbols denote BrS phenotype-positive, phenotype-negative, and phenotype-undetermined individuals, respectively. The pedigree has been modified to protect the privacy of family members; the changes do not affect the interpretation of segregation. Red and blue “+”/“−” symbols indicate carrier/non-carrier status for *SCN5A*:c.4883G>A (p.Arg1628Gln) and *KCND3*:c.1798G>A (p.Gly600Arg), respectively. CC, CT, and TT indicate rs12090194 genotypes (C, risk allele; T, protective allele).

Two variants were detected in 4/231 BrS cases from Thailand: c.1960G>A/p.Ala654Thr in three unrelated cases and c.1855A>G/p.Thr619Ala in one case. All four carriers were negative for BrS-associated rare and low-frequency *SCN5A* variants, i.e. rare coding variants, the Asian-specific *SCN5A*:p.Arg965Cys intermediate effect variant^12^, and the pathogenic non-coding variant in the RE5 enhancer element previously shown to be enriched in Thai BrS cases ^23^. Neither *KCND3* variant was detected in 500 ancestry-matched genome-sequenced controls or 1,092 Thai individuals in the Thai reference exome (T-REx) database ^24^. The p.Ala654Thr variant is rare in the general population - it has an allele frequency of 6.7e^−5^ in gnomAD v4.1-East Asian (EAS) samples and is absent from the Genomics Thailand database (n=14,466). The variant was therefore significantly enriched in BrS cases compared to 1,592 in-house controls (p=0.002), 22,452 individuals in gnomAD-EAS (p=2.0e^−5^), and 14,466 individuals in Genomics Thailand (p=3.8e^−6^). The p.Thr619Ala variant is absent from all gnomAD v4.1 populations but has an allele frequency of 0.000177 in the Genomics Thailand database (**Table 1**).

**Table 1.**
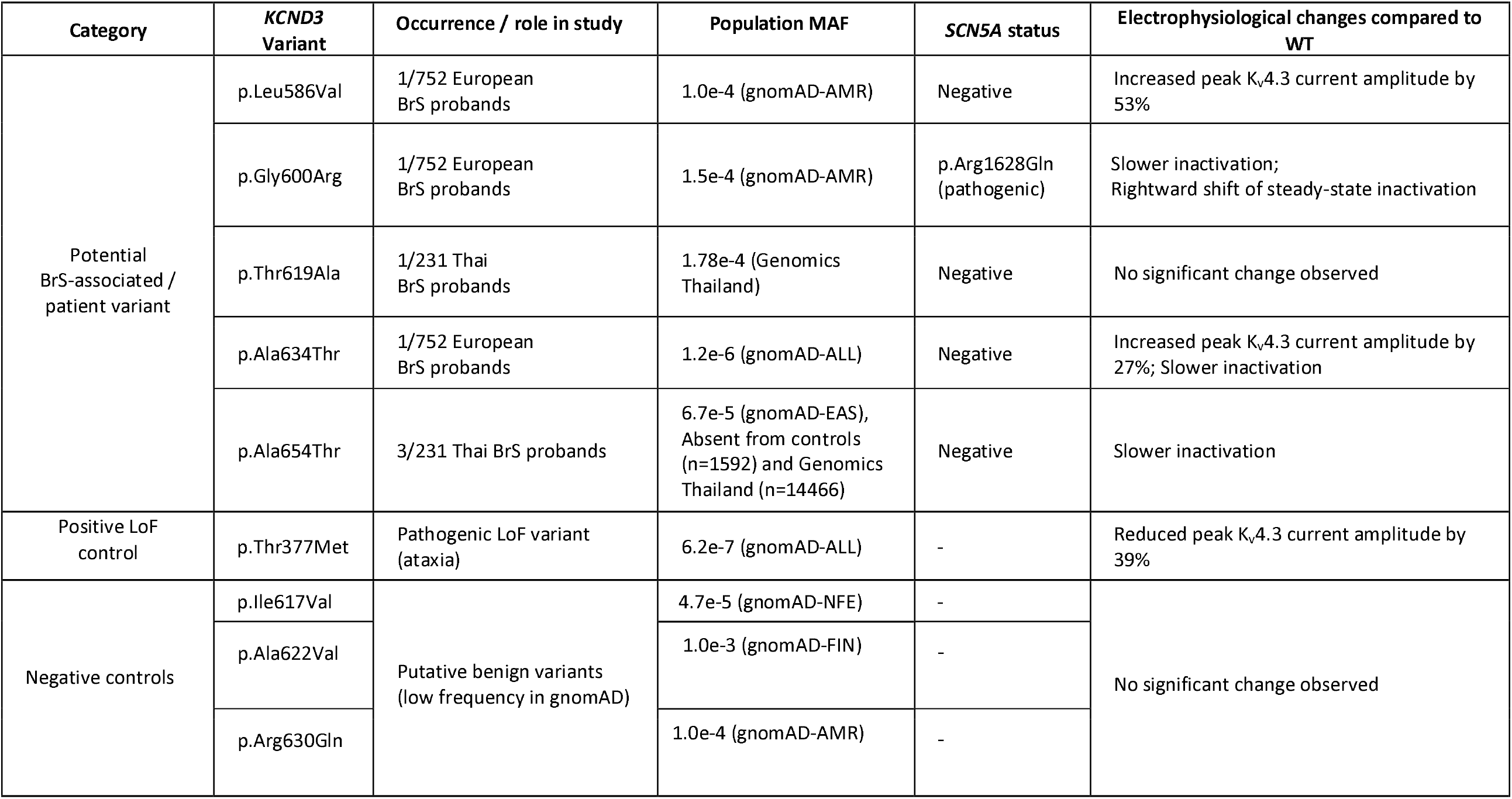
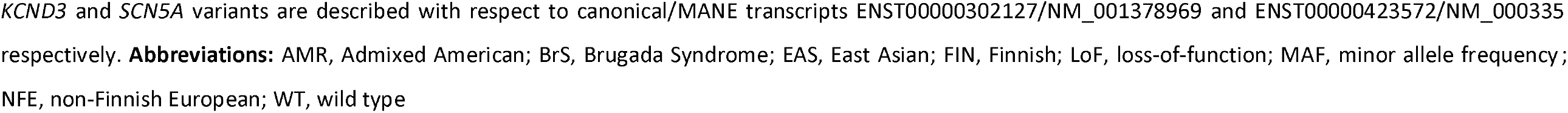
Genetic and functional summary of *KCND3* variants analysed in this study.

| Category | *KCND3* Variant | Occurrence / role in study | Population MAF | *SCN5A* status | Electrophysiological changes compared to WT |
| --- | --- | --- | --- | --- | --- |
| Potential BrS-associated / patient variant | p.Leu586Val | 1/752 European BrS probands | 1.0e-4 (gnomAD-AMR) | Negative | Increased peak K<sub>v</sub>4.3 current amplitude by 53% |
|  | p.Gly600Arg | 1/752 European BrS probands | 1.5e-4 (gnomAD-AMR) | p.Arg1628Gln (pathogenic) | Slower inactivation; Rightward shift of steady-state inactivation |
|  | p.Thr619Ala | 1/231 Thai BrS probands | 1.78e-4 (Genomics Thailand) | Negative | No significant change observed |
|  | p.Ala634Thr | 1/752 European BrS probands | 1.2e-6 (gnomAD-ALL) | Negative | Increased peak K<sub>v</sub>4.3 current amplitude by 27%; Slower inactivation |
|  | p.Ala654Thr | 3/231 Thai BrS probands | 6.7e-5 (gnomAD-EAS), Absent from controls (n=1592) and Genomics Thailand (n=14466) | Negative | Slower inactivation |
| Positive LoF control | p.Thr377Met | Pathogenic LoF variant (ataxia) | 6.2e-7 (gnomAD-ALL) | - | Reduced peak K<sub>v</sub>4.3 current amplitude by 39% |
| Negative controls | p.Ile617Val | Putative benign variants (low frequency in gnomAD) | 4.7e-5 (gnomAD-NFE) | - | No significant change observed |
|  | p.Ala622Val |  | 1.0e-3 (gnomAD-FIN) | - |  |
|  | p.Arg630Gln |  | 1.0e-4 (gnomAD-AMR) | - |  |

*KCND3* and *SCN5A* variants are described with respect to canonical/MANE transcripts ENST00000302127/NM\_001378969 and ENST00000423572/NM\_000335 respectively. **Abbreviations:** AMR, Admixed American; BrS, Brugada Syndrome; EAS, East Asian; FIN, Finnish; LoF, loss-of-function; MAF, minor allele frequency; NFE, non-Finnish European; WT, wild type

Four rare *KCND3* missense variants were detected in 752 BrS cases of European ancestry, each detected in one case and located in the C-terminal region of the protein. The c.1798G>A/p.Gly600Arg variant was found in a Dutch BrS patient who also carried the likely pathogenic *SCN5A*:p.Arg1628Gln variant (**Supplemental data**). p.Gly600Arg was previously described in a BrS patient ^17^ and an individual with sudden unexplained death ^25^. Notably, it was also detected in 1/1,827 controls in the present study and has a minor allele frequency of 0.0001 in gnomAD v4.1-non-Finnish European (NFE). The other three rare *KCND3* variants were each identified in a different French BrS case: c.1859C>A/p.Pro620His (this case also carried the pathogenic *SCN5A*:p.Arg893His variant, **Supplemental data**), c.1756C>G/p.Leu586Val and c.1900G>A/p.Ala634Thr (the latter two were sporadic *SCN5A*-negative cases) (**Table 1**).

### Electrophysiological K_v_4.3 changes in the variants

We next performed whole-cell patch-clamp recordings in transfected HEK293 cells to assess the electrophysiological impact of four of the novel K_v_4.3 variants identified in the BrS patients (p.Leu586Val, p.Thr619Ala, p.Ala634Thr and p.Ala654Thr; p.Pro620His was not prioritized as the proband’s affected son was a non-carrier), as well as the previously studied putative GoF variant p.Gly600Arg. For comparison, we also analysed three putatively benign variants (p.Ile617Val, p.Ala622Val, and p.Arg630Gln), which were prioritized for functional assessment based on their low allele frequencies in population databases and their localization in the same C-terminal region as the putative BrS variants. An established loss-of-function (LoF) variant associated with spinocerebellar ataxia (p.Thr377Met) was also included for comparison. The locations of all nine variants in the K_v_4.3 protein are illustrated in **Figure 1C**.

Patch-clamp recordings were performed on HEK293 cells transfected with wild-type (WT) and variant *KCND3* constructs at a 1:1 ratio to mimic the heterozygous state of the patients, together with the WT construct for *KCNIP2* encoding the K_v_4.3-modulating protein KChIP2. Recordings were performed in two experimental periods. As electrophysiological measurements are known to be sensitive to subtle changes in experimental conditions over time ^26^, each variant was compared with WT K_v_4.3 recordings from the same period, designated WT1 and WT2, respectively. All variants were compared with WT1, with the exception of p.Leu586Val and p.Ala634Thr, which were compared with WT2.

### Peak current amplitude is increased for *p*.*Leu586Val and p*.*Ala634Thr*

Representative K_v_4.3 current traces and quantification of peak current amplitudes at +40 mV are presented in **Figure 2A and 2B**. As expected, the p.Thr377Met variant exhibited a statistically significant 39% reduction in current amplitude (15.6 ± 1.9 nA (n=24) vs. 25.7 ± 1.8 nA (WT1; n=44)), consistent with its established LoF phenotype ^27^. Two variants, p.Leu586Val and p.Ala634Thr, displayed a significant 53% and 27% increase in K_v_4.3 current amplitude, respectively, compared to WT (35.9 ± 1.6 nA (p.Leu586Val; n=20), 29.8 ± 2.5 nA (p.Ala634Thr; n=24), and 23.5 ± 2.3 nA (WT2; n=26)) (**Figure 2B**), indicative of a GoF effect. Peak K_v_4.3 current amplitude did not significantly differ between WT, p.Thr619Ala and p.Ala654Thr, as well as the three putatively benign variants studied, suggesting that these variants do not impair macroscopic conductance. Although the peak current amplitude of the p.Gly600Arg variant showed a ≈25% increase compared to WT, this difference did not reach statistical significance (32.2 ± 2.2 nA (G600R; n=20) vs. 25.7 ± 1.8 nA (WT1; n=44)) (**Figure 2B**). Consistent with our findings, a 50% increase in current density (P<0.05) was observed for the p.Gly600Arg variant in a previous report that studied this variant under homozygous expression conditions ^17^.

**Figure 2.**
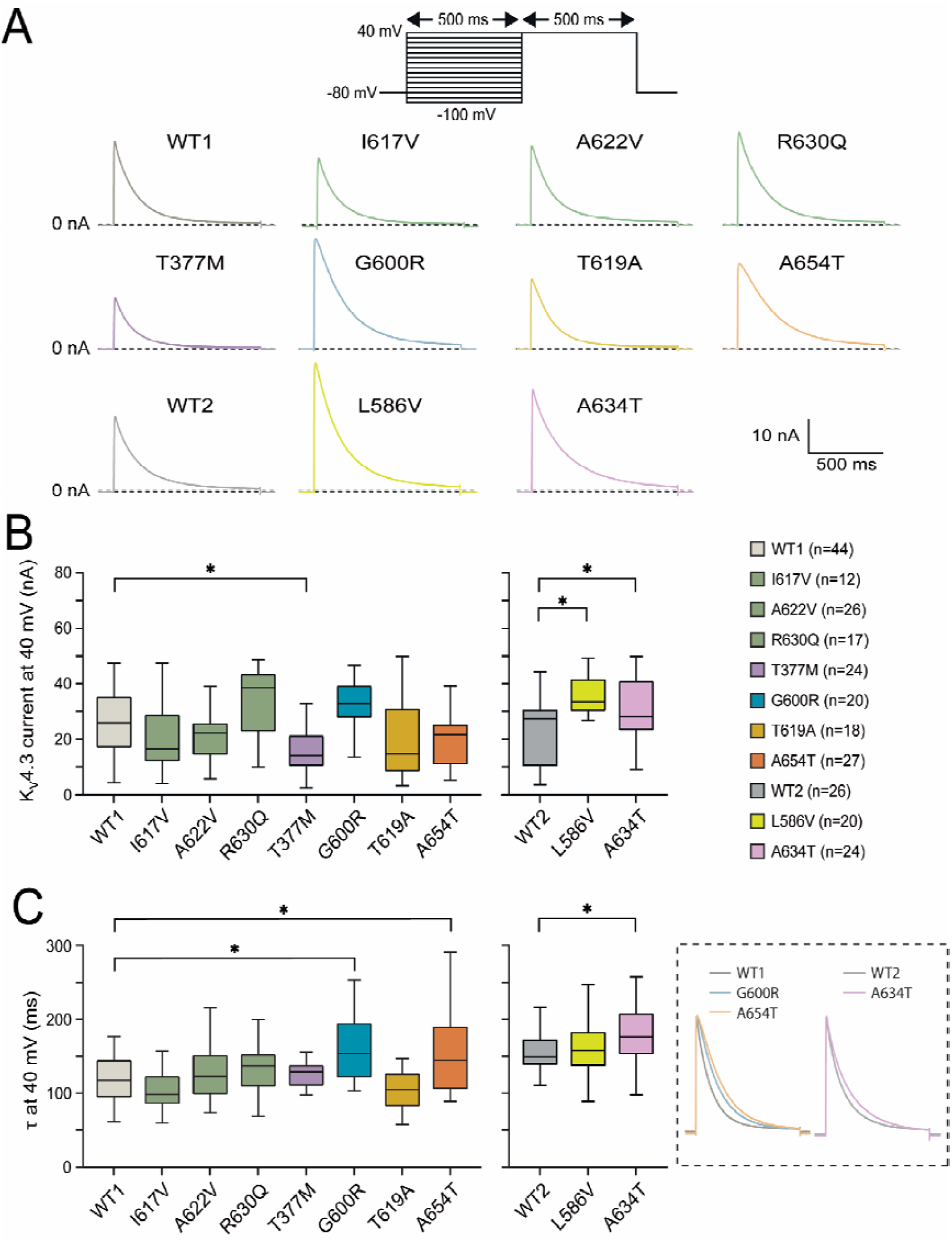
Effect of *KCND3* variants on K_v_4.3 current amplitude and decay time constant. **(A)** Representative current traces at a voltage clamp step from −80 to +40 mV from HEK293 cells co-expressing WT or mutant *KCND3* together with *KCNIP2*, encoding the putative K_v_4.3-modulating protein KChIP2. Recordings were obtained using a two-pulse voltage clamp protocol (shown at top). Comparator variants are color-coded throughout: the previously reported LoF variant T377M is shown in purple, whereas putatively benign comparator variants (I617V, A622V, and R630Q) are shown in dark green. **(B)** Quantification of peak K_v_4.3 current at +40 mV. Data are presented as box plots (median and boxes represent interquartile range), and variants were compared to WT currents recorded in the same patch clamp period (see Results section). L586V and A634T resulted in larger amplitudes, while the T377M amplitude was decreased. **(C)** Box plots showing the time constant of inactivation (*τ*) at +40 mV for WT and all tested *KCND3* variants. Inset: Representative current traces illustrating the slower decay of current in the G600R, A654T, and A634T variants. *p < 0.05, one-way ANOVA followed by Holm–Šidák post hoc test.

### Slower inactivation kinetics for the p.Gly600Arg, p.Ala634Thr and p.Ala654Thr variants

As shown in **Figure 2C**, inactivation kinetics analysis at +40 mV revealed a significant increase of the inactivation time constant (*τ*) for the BrS-associated p.Gly600Arg, p.Ala654Thr and p.Ala634Thr variants compared to WT measured in the corresponding period (p.Gly600Arg, 161.0 ± 10.0 ms (n=20), p.Ala654Thr, 155.2 ± 11.3 ms (n=27), versus WT1, 120.3 ± 4.6 ms (n=44); p.Ala634Thr, 179.4 ± 7.9 ms (n=24) versus WT2, 151.7 ± 6.1 ms (n=26)). No significant changes were observed in all other variants studied. Notably, the slower inactivation kinetics observed for p.Gly600Arg are consistent with previous findings ^17^. The slower inactivation displayed by these three variants prolongs channel opening before it transitions to the non-conducting state and is expected to lead to an increase in net outward current.

### Minor changes in voltage dependence of (in)activation

The voltage-dependent gating properties of all K_v_4.3 variants tested are summarized in **Supplemental Figures 1 and 2**. All variants exhibited half-activation and half-inactivation voltages (*V*_*1/2*_) similar to WT, except for the BrS-associated p.Gly600Arg variant, which had a significant rightward shift of 6.6 mV in steady-state inactivation, as illustrated in **Supplemental Figure 2B (inset)** (*V*_*1/2*_ inactivation: −32.6 ± 1.3 mV vs. WT1 −39.2 ± 0.8 mV). This shift indicates that the K_v_4.3 channel requires a less negative membrane potential to reach the same level of inactivation in the presence of p.Gly600Arg (there will be less inactivation at each membrane potential). Subtle, but statistically significant alterations in the slope factor (*K*) of the voltage dependence of channel opening were observed for the p.Ala622Val and p.Ala654Thr variants. Both variants exhibited an increased activation slope factor (*K*: 11.4 ± 0.3 and 11.8 ± 0.3 mV, respectively, vs. WT1 10.6 ± 0.3 mV). However, the *K* value changes are limited while the voltage range of activation is passed quickly during the very fast action potential upstroke, so the observed changes in the *K* of activation will have limited functional consequences.

### Clinical characteristics of patients with KCND3 variants

We explored the clinical characteristics of the cases carrying functionally validated GoF variants in *KCND3* (ECGs of the respective patients are presented in **Supplemental Figure 3**). All three Thai BrS patients carrying the p.Ala654Thr variant are male, symptomatic and have a family history of SCD. Patient 1 experienced his first cardiac event in his twenties with ventricular tachycardia/ventricular fibrillation (VT/VF). A type 1 Brugada ECG pattern was induced during sodium channel blocker challenge (**Supplemental Figure 3A**). Patient 2 presented with VT/VF arrest in his forties, with a post-arrest ECG demonstrating a spontaneous type 1 Brugada ECG pattern (**Supplemental Figure 3B**). His father’s sibling was reported to have died of SCD (age and genotype unknown). Patient 3 suffered cardiac arrest in his thirties, with electrophysiological study revealing inducible polymorphic ventricular tachycardia (PMVT)/VF, and a spontaneous type 1 Brugada ECG pattern was observed (**Supplemental Figure 3C**). The patient’s brother died of SCD in his twenties (genotype unknown).

The patient carrying the p.Ala634Thr variant was male and diagnosed with BrS in his thirties. He had an intermittent spontaneous type 1 Brugada ECG pattern (**Supplemental Figure 3D**) but no inducible PMVT/VF during an electrophysiological study. He had no family history of BrS or SCD and remained asymptomatic during follow-up. The p.Leu586Val carrier was male and diagnosed with BrS in his forties based on a drug-induced BrS pattern (**Supplemental Figure 3E**). His father died of SCD in his forties (genotype unknown). The patient remained asymptomatic until he died due to a non-cardiac cause in his fifties.

The patient carrying the p.Gly600Arg variant (and the likely pathogenic *SCN5A*:p.Arg1628Gln variant) was a male diagnosed during occupational screening. Baseline ECG showed a type 2 Brugada ECG pattern. Exercise testing revealed a wide complex tachycardia with left axis deviation and right bundle branch block (RBBB) morphology, which was followed by sinus rhythm with coved ST segment elevation of 5mm during recovery phase. Subsequently, a PMVT was induced during an electrophysiological study. An implantable cardioverter-defibrillator (ICD) was implanted and spontaneous type 1 Brugada ECG pattern was observed repeatedly during follow-up (**Supplemental Figure 3F**), although he remained free of major symptoms. Analysis of his family pedigree revealed two other carriers of both variants (affected brother, unaffected sister) and two carriers of the p.Gly600Arg variant only - his sister had a positive ajmaline test and vasovagal syncope, and his daughter, in her twenties, displayed incomplete RBBB with no ST elevation (ajmaline testing not performed) (**Figure 1D, Table S3**).

To identify additional carriers of these BrS-associated variants, we searched the genetic testing results for over 40,000 individuals referred for clinical genetic testing for a range of inherited cardiac conditions at Health in Code, Spain. One patient carried the p.Ala654Thr variant - a woman from Spain in her forties who was referred for genetic testing and whose brother had died suddenly in his sleep while in his twenties (genotype unknown). She presented with dizziness and a syncopal episode with a clear vagal profile, had a normal cardiac MRI (no evidence of late gadolinium enhancement), no ECG abnormalities detected either at baseline or after flecainide and ajmaline provocation, and no recorded events on an implantable loop recorder over the course of a year despite episodes of dizziness and presyncope. There were no cases with the p.Ala634Thr variant, while p.Leu586Val and p.Gly600Arg were detected at background population frequencies in non-BrS referrals.

### Potential epistatic effects of BrS-associated common variant at KCND3 locus

The GWAS signal at the *KCND3* locus that was significantly associated with BrS comprises a group of several common variants in high linkage disequilibrium located within the large first intron of *KCND3* (**Figure 3A** and **3B**). Expression quantitative trait locus (eQTL) data from the GTEx portal ^28^ demonstrate that the risk allele for BrS at the lead GWAS SNP (C, the reference and major allele of rs12090194) is associated with increased expression of *KCND3*, consistent with the proposed GoF mechanism for K_v_4.3/*I*_*to*_ in the disorder (**Figure 3C**). We hypothesized that direct epistatic interaction between this common variant and the rare GoF *KCND3* missense variants could influence disease penetrance in carriers of these pathogenic variants ^29^.

**Figure 3:**
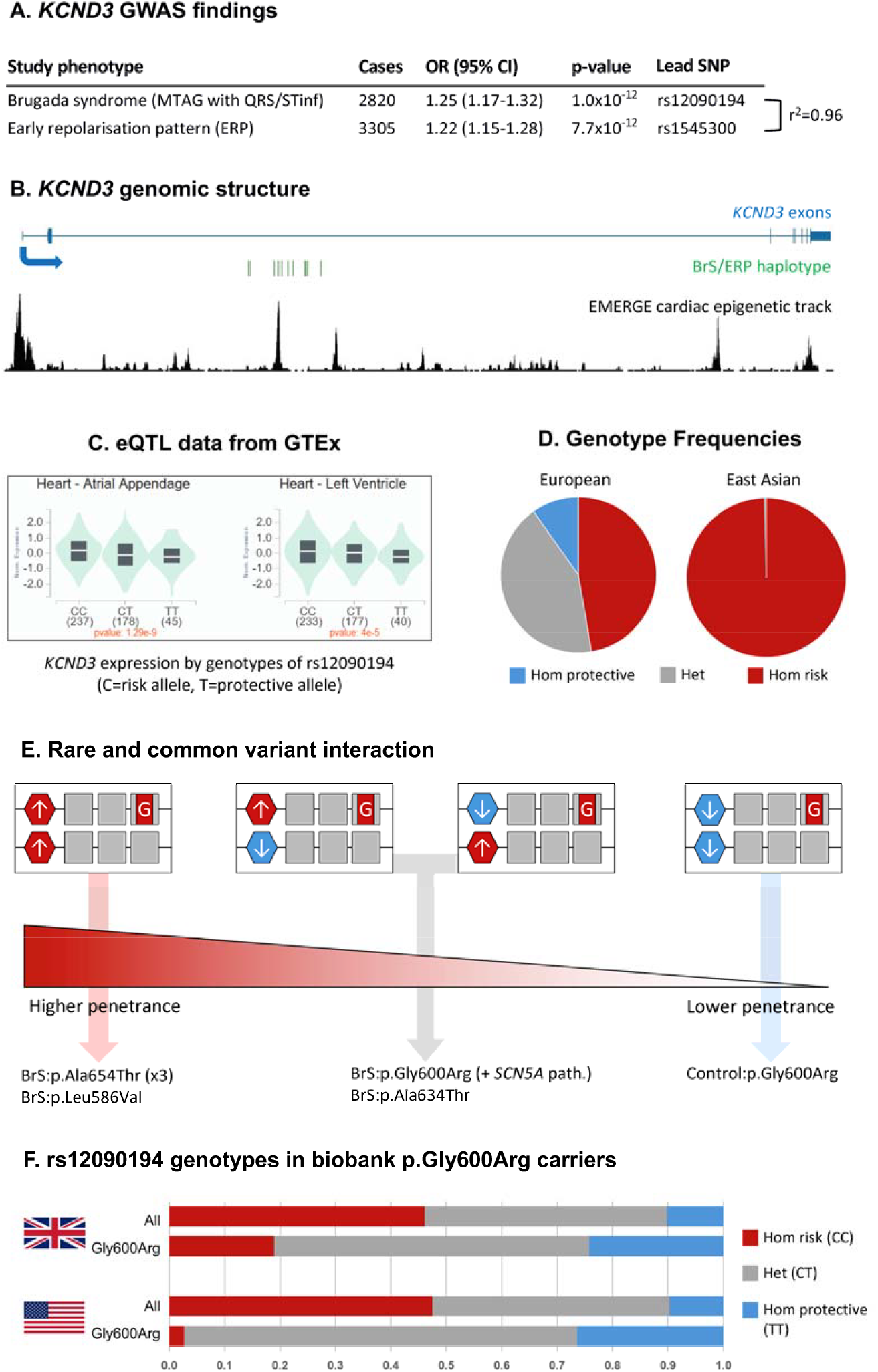
Potential interactions between rare gain-of-function (GoF) coding variants in *KCND3* and common risk variants at this locus. **(A)** Common variants at the *KCND3* locus have been identified in two key genome-wide association studies (GWAS) – a Brugada Syndrome (BrS) case-control study utilising multi-trait analysis of GWAS (MTAG), incorporating the correlated ECG endophenotype traits of QRS duration and ST inferior (STinf), and a GWAS on the related J-wave phenotype of early repolarization pattern (ERP). The lead SNPs in both studies are in strong linkage disequilibrium (LD) with each other (r^2^=0.96). **(B)** These two lead GWAS SNPs, and 12 other common variants in close LD (r^2^>0.8) are located within the large (194.5kb) intron 2 of *KCND3. KCND3* exons are highlighted by the blue bars (taller bars for coding sequence, shorter bars for non-coding). The EMERGE track of cardiac-specific epigenetic markers is shown in black^49^. **(C)** Expression quantitative trait locus (eQTL) from GTEx demonstrates that these SNPs are associated with altered expression of *KCND3* in heart tissues, with the risk allele (C) associated with increased expression, consistent with the GoF effect of K_v_4.3 in BrS (GTEx Portal, accessed 31/08/2026). **(D)** The risk allele is the major allele in the population, particularly in East Asia where >99% of individuals carry two copies. **(E)** Interaction between these common risk alleles and rare coding GoF variants may partly influence penetrance in rare variant carriers. Common alleles are indicated by hexagons (C risk allele in red, T protective allele in blue) and rare GoF variants indicated by red squares (G). Increased expression of the mutated allele, and to a lesser extent the wild type allele, may make it more likely that an individual develops the BrS phenotype, as this may contribute to the GoF effect. The three BrS carriers of *KCND3*:p.Ala654Thr and one carrier of *KCND3*:p.Leu586Val were all homozygous for the risk allele while the control carrier of p.Gly600Arg was homozygous for the protective allele (a genotype occurring in <10% of Europeans). **(F)** Comparison of the rs12090194 genotypes in carriers of the *KCND3*:p.Gly600Arg in the UK Biobank (top) and *All of Us* (bottom) with all non-Finnish European individuals, which demonstrates that the risk C allele is significantly depleted in these rare variant carriers (based on population frequency, most of these carriers are expected to be unaffected).

The common risk allele at this locus (rs12090194-C) has a frequency of 0.998 in gnomAD East Asian samples and occurs in the homozygous state in the three p.Ala654Thr BrS cases in this study, indicating increased expression of both WT and mutated alleles in these patients, which may lead to a more pronounced GoF effect and therefore expression of the BrS phenotype. The alternate protective allele (rs12090194-T) is present at a higher frequency in non-East Asian populations, with an allele frequency of 0.313 in gnomAD-NFE. Therefore, 9.8% of Europeans are homozygous for the protective allele (associated with reduced *KCND3* expression) and 42.9% have a heterozygous genotype. In contrast, >99% of East Asians are homozygous for the risk allele (**Figure 3D**).

For the European BrS patients with *KCND3* GoF variants described here, the patient carrying p.Leu586Val is homozygous for the risk rs12090194 C allele, while the p.Gly600Arg and p.Ala634Thr carriers are heterozygous. Phasing of the common and rare variants could not be resolved for the p.Ala634Thr carrier, but pedigree analysis of the p.Gly600Arg family indicates that the rare variant co-occurs with the protective T allele in this family. However, as this patient also harbours a pathogenic *SCN5A* coding variant, epistatic effects at the *KCND3* locus are unlikely to substantially influence phenotype expression. In contrast, the control p.Gly600Arg carrier was homozygous for the protective allele (**Figure 3E**). To further explore potential epistatic effects in these carriers, we investigated the common variant genotypes in 95 European-ancestry p.Gly600Arg carriers in the UK Biobank. The frequency of the protective allele (rs12090194-T) was significantly enriched in these individuals (0.526 compared to 0.32 in all UK Biobank NFE individuals, p=5.1e^−9^). Notably, 24.2% of the carriers were homozygous for the protective allele, a 2.4-fold enrichment compared to the population. These findings were replicated in 38 European-ancestry p.Gly600Arg carriers in *All of Us*, where the rs12090194-T allele had a frequency of 0.618 (compared to 0.311 for all NFE individuals, p=3.6e^−8^) and 26.3% of carriers were homozygous for the protective allele (compared to 9.7% for all NFE individuals) (**Figure 3F**) (**Table S4**). This may offer at least a partial explanation for the relatively high population frequency of this proven GoF variant (1.1×10^−4^ in NFE from gnomAD v4.1.1), which may reflect linkage disequilibrium between rs12090194 and Gly600Arg on a shared ancestral haplotype.

## Discussion

We here used genome sequencing data from a multi-ancestry cohort of 983 BrS probands from Europe and Thailand to explore the role of *KCND3* variants in BrS, identifying an enrichment of rare C-terminal variants in cases and demonstrating GoF effects for four variants using functional assays. Two variants (p.Leu586Val and p.Ala634Thr) increased peak current amplitude, while two others (p.Ala654Thr and p.Gly600Arg) slowed K_v_4.3 inactivation without increasing peak current amplitude; the p.Gly600Arg variant also shifted steady-state inactivation to less negative potentials, consistent with increased channel availability. Such GoF effects are concordant with both the prevailing hypothesis for BrS pathomechanisms and GWAS findings at the *KCND3* locus for BrS.

BrS has proven to be one of the most intractable cardiac genetic disorders with regard to elucidating its genetic aetiology. Its complex genetic nature is highlighted by an unexpectedly large polygenic component revealed by GWAS and limitations in familial genetic analysis, which typically involve modestly-sized pedigrees and even occasionally demonstrate non-segregation of pathogenic *SCN5A* variants (i.e. individuals lacking the primary causal familial variant can still present with a BrS phenotype) ^30,31^. Although approximately 30 genes have been suggested to harbour causal rare variants for BrS, *SCN5A* remains the only clinically actionable gene almost 30 years since its discovery ^19^. It remains unclear to which extent the ClinGen conclusions for BrS (all genes except *SCN5A* were classified as disputed) reflect invalid associations or rather an inability to account for its complex genetic architecture, as BrS-associated genes and variants may not conform to the paradigms of a classical Mendelian disorder. Putative novel genetic risk factors may not segregate strongly with phenotype in families, display limited or nuanced case-control enrichment, or have modest effect sizes and consequently not ultra-rare in population databases. To overcome these challenges, we require novel approaches and the judicious appraisal of multiple orthogonal lines of evidence for identifying novel BrS genes, that maintain stringent contemporary standards for defining genetic associations while not necessarily being bound by Mendelian conventions that may not fully apply to BrS.

We believe the data presented here across orthogonal classes of evidence establishes *KCND3* as a BrS disease gene. This includes robust genetic evidence for both common and rare variation - a genome-wide significant locus at *KCND3* and case enrichment of rare missense variants in the K_v_4.3 C-terminus, respectively. While GWAS signals are not included as evidence for ClinGen gene-disease curation, convergence between rare and common variant associations is widely observed at disease loci, including the *SCN5A* locus in BrS ^9–13^ and key loci in other cardiac genetic diseases like long QT syndrome ^32^ and hypertrophic cardiomyopathy ^33^. We demonstrate GoF effects for four of the rare C-terminal variants found in cases through functional experiments that importantly incorporate both rare control variants and heterozygous co-expression of WT and variant *KCND3* constructs to reflect the patient genotypes. Additionally, our data highlight a possible epistatic common-rare variant interaction at this locus, with a significant enrichment of the protective expression-reducing allele observed in carriers of the p.Gly600Arg variant across two population biobanks (based on the variant frequency most of these carriers will be unaffected). This interaction may modify penetrance in carriers of rare GoF variants and partially explain prior conflicting observations between functional and population genetic data. Taken together, this demonstrates the complex mechanisms by which GoF of *KCND3*/K_v_4.3 contributes to the pathophysiology of BrS.

Many genes have been implicated in BrS and subsequently disputed despite published studies demonstrating a functional effect of variants detected in cases. Rare missense variants are difficult to evaluate in isolation, particularly in heterologous expression systems where small differences may reflect experimental variability rather than true variant-specific dysfunction. To address this, an important aspect of our study design was the inclusion of comparator and reference rare *KCND3* variants to contextualize the results. We assessed three rare, putatively benign variants located in the same C-terminal region as the case variants that provided a local functional baseline for rare variation in this region. Against this background, we were able to distinguish between the four GoF variants and the case variant p.Thr619Ala which showed no clear deviation from the benign comparator variants, supporting its interpretation as functionally neutral under these experimental conditions. In parallel, p.Gly600Arg and p.Thr377Met served as established functional references, confirming that the assay could detect both BrS-relevant GoF and ataxia-associated LoF effects, respectively. Together, this design provided an internally calibrated framework for distinguishing functionally relevant *KCND3* variant effects from background variability. In our more conservative co-expression system, where variant constructs were co-expressed with WT to mimic the heterozygous genotype in the patients, p.Gly600Arg showed only a modest, non-significant increase in peak K_v_4.3 current amplitude compared with WT, in contrast to the larger current increase reported under mutant-only conditions ^17^. Nevertheless, key GoF features remained evident, including delayed inactivation and a depolarizing shift in steady-state inactivation, consistent with increased channel availability.

A gain-of-function in the transient outward current *I*_*to*_, in humans mediated primarily by the K_v_4.3 channel encoded by *KCND3*, has long been hypothesized to contribute to the BrS pathogenesis. In one proposed mechanism (the repolarization hypothesis ^14^), an increase in *I*_to_ leads to accentuation of transmural and regional differences in cardiomyocyte phase 1 repolarization. Such differences potentially lead to the loss of the action potential dome and marked regional variations in action potential duration, with steep repolarization gradients leading to phase 2 reentrant extrasystoles that may cause ventricular fibrillation ^4,34^. In a second proposed mechanism, increased *I*_to_ may contribute to delayed depolarization by opposing inward depolarizing currents, thereby reducing the conduction safety factor and promoting activation slowing and conduction block, particularly within the vulnerable right ventricular epicardium in the presence of microstructural abnormalities ^15,35^. This GoF model for *I*_*to*_ in BrS and our experimental findings, align with the clinical efficacy observed for the Class Ia antiarrhythmic drug quinidine - which blocks *I*_to_ - in patients with the disorder ^36,37^. Of note, the GWAS-identified BrS risk locus at *KCND3* (tagged by rs12090194) has also been associated with early repolarization pattern, an endophenotype of early repolarization syndrome, in population-based GWAS ^38^, supporting the contemporary view that these disorders are mechanistically related (J wave syndromes) ^3,4^. In line with this, rare *KCND3* variants have also been reported in isolated cases of early repolarization syndrome, including a *de novo* GoF variant ^39^ and large structural duplication including *KCND3*^40^.

The clustering of BrS-associated *KCND3* variants within the distal C-terminal region is unlikely to be incidental and possibly provides a mechanistic context for the functional effects observed in this study. A number of studies that generated truncating mutations in the K_v_4.3 C-terminal domain, including the distal region, reported effects on K_v_4.3 functional expression and gating properties^41,42^. The C-terminal region is the binding location for a number of K_v_4.3 interacting proteins, including the canonical modulatory subunit KChIP2, which increases *I*_*to*_ amplitude and modifies inactivation and recovery kinetics^43^. Functional studies have shown that progressive C-terminal truncation attenuated KChIP2-dependent modulation of K_v_4.3, and suggested that the region distal to residue 486 may contribute, directly or indirectly, to this regulation^42^. Furthermore, two proline-rich sequences in K_v_4.3 bearing the PXXP motif, at residues 618–621 and 630–636, were proposed as potential interaction sites for the SH3 domain of the tyrosine kinase c-Src, the activity of which enhances K_v_ 4.3 current amplitude ^44^. Notably, the p.Pro620His and p.Ala634Thr variants identified in BrS patients in this study are located within these PXXP sequences. Finally, the terminal SAL residues of K_v_4.3 have been shown to interact with the scaffolding protein SAP97, which promotes K_v_4.3 clustering at the plasma membrane, increases K_v_4.3 current density and supports CaMKII-dependent channel regulation ^45^. Of particular relevance, the p.Ala654Thr variant we identified alters the central residue of this motif. Together, these observations raise the possibility that altered protein interactions at the distal K_v_4.3 C-terminus may contribute to the functional effects of these variants. While the cryo-electron microscopy structure of the K_v_4.3–KChIP2 complex demonstrated C-terminal contacts involving the proximal region adjacent to S6^46^, the distal K_v_4.3 C-terminal tail was poorly resolved.

Based on the findings of this study, we believe the inclusion of *KCND3* in BrS clinical genetic testing is warranted, although the interpretation of findings may be challenging. Any novel missense variant detected in BrS patients, especially one located in the C-terminus, will require functional validation, particularly in the absence of robust genetic evidence (such as confirmed de novo inheritance). As this may be onerous for most laboratories, *KCND3* could be prioritized for high throughput functional mapping approaches (such as saturation mutagenesis coupled to high-throughput patch-clamp) that can proactively identify all potential GoF missense variants in this gene or its C-terminal region, as done previously for other ion channel subunits ^47,48^. Genotyping of the common rs12090194 risk allele may also be informative in estimating disease risk and penetrance for patients and their family members. Additionally, as we begin to evaluate the clinical utility of polygenic scores for diseases like BrS, potential epistatic interactions in carriers of rare GoF *KCND3* variants may need to be incorporated beyond the simple additive summation of common risk variants.

We acknowledge some limitations in this study. While the HEK293 expression system provides a controlled system for isolating variant-specific effects on K_v_4.3 function, it does not recapitulate the full cardiomyocyte environment in which native *I*_*to*_ operates and may miss certain components of the cardiac K_v_4.3 channel complex. Patch-clamp recordings were performed at room temperature and under ionic conditions optimized for K_v_4.3 current measurement, which differ from the physiological conditions present in human cardiomyocytes. The effects of the *KCND3* GoF variants are likely to be influenced by a broader array of background genetic variation, which may include additional variants at the *KCND3* locus and the overall BrS polygenic score.

In conclusion, we present detailed genetic and functional evidence to establish a critical role for *KCND3*/K_v_4.3 in the pathogenesis of BrS, resolving the prior uncertainty of this gene-disease association. Future studies are required to explore the broader effects of K_v_4.3 GoF in multicellular cardiomyocyte models, including whether these variants influence conduction reserve, repolarization heterogeneity, or arrhythmia susceptibility. It will also be critical to comprehensively evaluate how GoF variants interact with background genetic variation at both the *KCND3* locus and genome-wide, to inform future genetic risk profiling scores for enhancing diagnosis and risk prediction in BrS.

## Data Availability

All data produced in the present study are available upon reasonable request to the authors

## Acknowledgements

We thank the biological resource centre for biobanking (CHU Nantes, Nantes Université, Centre de resources). We are grateful to the Genomics Core Facility GenoA, member of Biogenouest and France Genomique, and to the Bioinformatics Core Facility BiRD, member of Biogenouest and Institut Français de Bioinformatique (IFB) (ANR-11-INBS-0013) for the use of their resources and their technical support. We are grateful to the support of the GenOmic variability in heaLth & Disease “GOLD”, a national transversal research project supported by INSERM. The authors thank all the participants and professionals contributing to the UK Biobank. All analyses for this study were conducted under UK Biobank application number 742. We gratefully acknowledge *All of Us* participants for their contributions, without whom this research would not have been possible. We also thank the National Institutes of Health’s *All of Us* Research Program for making available the participant data examined in this study. This study used data from the *All of Us* Research Program’s Controlled Tier Dataset [version 8], available to authorised users on the Researcher Workbench (https://workbench.researchallofus.org/). Access to the *All of Us* registered and controlled data tiers was granted through the Researcher Workbench, with City St. George’s (University of London) being approved under a Data Use and Registration Agreement.

## Data availability statement

The data that support the findings of this study are available from the corresponding author upon reasonable request.

