## Supplemental materials for "Convergent genetic and functional evidence implicate *KCND3* in Brugada Syndrome"

#### Supplemental methods

##### Patient cohorts

The BrS cohorts in this study comprised two groups - unrelated cases of European-ancestry (n=752) and unrelated cases from Thailand (n=231). The European cohort was recruited from Amsterdam University Medical Centre and Maastricht UMC+ in the Netherlands (n=262), through cardiology departments of university hospitals participating to the national healthcare network for inherited cardiac diseases in France (Cardiogen; in this study specifically the university hospitals in Nantes, Brest and Bordeaux) (n=343) and St George's University Hospitals NHS Foundation Trust and University College London in the UK (n=147). Patients in the Thai cohort were recruited from 7 participating institutions in Thailand.

All patients were diagnosed with BrS according to the 2013 Heart Rhythm Society/European Heart Rhythm Association/Asia Pacific Heart Rhythm Society Expert consensus statement [1], the 2015 guidelines of the European Society of Cardiology [2] and the 2017 guidelines of the American Heart Association[3]. All cases had ≥2 mm type I ST-segment elevation in ≥1 right precordial lead (V1–V2) of standard 12-lead ECG, either spontaneously or after provocation using a class I antiarrhythmic drug administered intravenously. ECGs were centrally reviewed by a BrS-expert electrophysiologist.

UK and Dutch controls were obtained from Project MinE [4] and French controls were obtained from the FranceGenRef consortium (n=1827 total European ancestry controls). Thai controls (n=500) were local volunteers of Thai ethnic origin by self-report without type 1 Brugada pattern on standard 12-lead ECG. As Southeast Asian individuals are poorly represented in population databases like gnomAD; two additional national genomics resources were utilized to assess variant rarity - the Thai reference exome (T-REx) variant database (n=1092) [5] and the Genomics Thailand database (n=14,466).

The study was approved by the relevant ethics committees in the Netherlands (Amsterdam UMC METC: W20_226 #20.260), France (Nantes: DC-2011-1399), the UK (St George's: REC Ref: 10/H0803/121, IRAS number 53663; UCL: 15/LO/0549) and Thailand (Chulalongkorn University Faculty of Medicine institutional review board no. 431/58). Informed consent was obtained from all participants.

#### Family cascade screening

Detailed clinical information was retrieved for probands harbouring rare *KCND3* variants. Where available, we collected clinical data of relatives and conducted cascade genetic testing for co-segregation analysis and phasing of rare and common variants.

#### Genome Sequencing

Short-read genome sequencing data was generated for the cases and controls using Illumina HiSeqX with a PCR-free library. Data processing, alignment, and variant calling were performed using standardised approaches as previously described [6,7]. Samples were excluded due to: high missing genotype rate (>0.03), relatedness (PI_HAT>0.1; one of related sample group retained with priority for cases), high heterozygosity rate (>0.4), high inbreeding rate (F>0.1), or sex check inconsistencies. Sample ancestry was determined using principal component analysis (PCA) with samples from the 1000 Genomes (1KG) data set and any samples not clustering with northwest European or East Asian samples for the respective datasets were excluded.

#### Genetic analysis of *KCND3*

Rare missense and inframe insertion/deletion variants in *KCND3* were identified in the case and control cohorts, with rarity defined as a gnomAD v4.1 filtering allele frequency (FAF) <0.0001 (using the joint exome/genome metric). All variants are described with respect to the *KCND3* canonical/MANE transcript ENST00000302127/NM_001378969. For cases with rare *KCND3* variants, their *SCN5A* genetic status was assessed by noting rare protein-altering variants or previously described *SCN5A* risk factors in Asian populations [6]. *SCN5A* variants are described with respect to the canonical/MANE transcript ENST00000423572/NM_000335 (2015 amino acids). Rare variant case-control enrichment was tested by comparing case frequencies with both the study controls and gnomAD v4.1 exomes datasets, using all gnomAD samples (for European and Thai cases combined) and non-Finnish European (NFE) and East Asian (EAS) samples for ancestry-specific comparison. Analysis was performed across the entire *KCND3* gene/transcript and separately for the final exon where case variants were clustered, with statistical significance assessed using Fisher's exact test.

The genotype for the common BrS risk variant rs12090194 was noted in cases and controls carrying rare *KCND3* missense variants. Individuals of NFE ancestry in the UK Biobank and *All of Us* datasets that carried the *KCND3*:p.Gly600Arg variant were identified and their rs12090194 genotype was extracted. rs12090194 allele frequencies and genotypes for p.Gly600Arg carriers were compared to all NFE individuals for each biobank dataset. UK Biobank analyses for this study were conducted under application number 742. Access to the *All of Us* registered and controlled data tiers was granted through the Researcher Workbench, with City St. George's (University of London) being approved under a Data Use and Registration Agreement.

#### Plasmid mutagenesis

The pIRES2-*KCND3*$^{WT}$-EGFP and pIRES2-*KCNIP2*$^{WT}$-dsRed2 plasmids were generously provided by Dr. Michael Ackerman's laboratory [8]. Variant *KCND3* constructs were generated using site-directed mutagenesis, with the pIRES2-*KCND3*$^{WT}$-EGFP plasmid serving as the template. Mutagenesis was performed using the QuickChange XL Mutagenesis Kit (Agilent Technologies) in conjunction with custom-designed primers for specific nucleotide substitutions. Following mutagenesis, whole-plasmid sequencing was conducted to verify the successful introduction of each variant and to confirm the overall integrity of the constructs, ensuring the absence of unintended mutations.

#### HEK293 cell culture and transfection

HEK293 cells constitutively overexpressing the human $Na_v1.5$ channel [9] were cultured in DMEM (Dulbecco's Modified Eagle Medium, Gibco) supplemented with 10% fetal bovine serum (Biowest), 1% penicillin-streptomycin (Gibco), and 200 μg/mL Zeocin (Thermo Fisher Scientific) at 37°C in a 5% $CO_2$ incubator. Cells were transfected at approximately 60% confluency in 6-well plates with 20 ng total *KCND3* plasmid DNA per well, consisting of either wild-type (WT) *KCND3* alone or WT and variant *KCND3* constructs at a 1:1 ratio to mimic the heterozygous genotype of patients, together with 200 ng *KCNIP2*$^{WT}$ construct. Transfection was done using 0.9 µL of Lipofectamine 2000 (Thermo Fisher Scientific) in DMEM. At 48h post-transfection, cells were trypsinized and kept at room temperature (21°C) in complete DMEM until patch-clamp recording.

#### Patch-clamp analysis of $K_v4.3$ current

For patch-clamp experiments, cells were transferred to a recording chamber mounted on the stage of an inverted microscope (Nikon Eclipse TE2000-U), and single cells co-expressing green and red fluorescence were selected for recordings. Currents generated by $K_v4.3$ in the presence of KChIP2, hereafter referred to as $K_v4.3$ currents, were measured in the whole-cell configuration of the patch-clamp technique using an Axopatch 200B amplifier (Molecular Devices). Voltage control, data acquisition, and analysis were accomplished using custom software. Signals were low-pass filtered with a cut-off frequency of 5 kHz and digitized at 4 kHz. Patch pipettes were pulled from borosilicate glass (TW100F-3, World Precision Instruments Germany GmbH) and had resistances of ≈2 MΩ after filling with the internal solution described below. Series resistance was compensated by ≥80%. Cell membrane capacitance ($C_m$) was calculated by dividing the time constant of the decay of the capacitive transient upon 5 mV hyperpolarizing voltage step from −40 mV by the series resistance.

The $K_v4.3$ currents were relatively large and therefore, experiments were performed at room temperature using an elevated extracellular $K^+$ concentration to reduce the driving force ($E_K$). The external bath solution contained (in mM): 125.4 NaCl, 20 KCl, 1.8 $CaCl_2$, 1 $MgCl_2$, 5.5 glucose, and 5 HEPES (pH 7.4, adjusted with NaOH). Patch pipettes were filled with solution containing (in mM): 125 K-gluconate, 20 KCl, 5 NaCl, 1 $MgCl_2$, 10 EGTA, 5 MgATP, and 10 HEPES (pH 7.2, adjusted with KOH). A two-pulse protocol was employed to assess $K_v$4.3 current amplitude and gating properties, as illustrated in **Figure 2A**, inset. For each voltage step, peak $K_v$4.3 current amplitudes were analysed. Since the peak current amplitudes were not correlated with $C_m$, current amplitudes are expressed in nA. For analysis of the voltage dependency of (in)activation, currents were normalized to the maximal current amplitude; activation curves were additionally corrected for the electrochemical driving force. Steady-state (in)activation curves were fitted using the Boltzmann equation: $I / I_{max} = 1 / [1 + \exp((V_{1/2} - V) / K)]$, where $I / I_{max}$ is the normalized current, $V$ is the membrane potential, $V_{1/2}$ is the half-activation or half-inactivation voltage, and $K$ is the slope factor. The decay time constant ($\tau$) at +40 mV was obtained by fitting the inactivating phase of the current to a mono-exponential function using the following equation: $I_{(t)} = A \times \exp(- t / \tau) + C$, where $I_{(t)}$ is the current at time $t$, $A$ is the amplitude of the time-dependent component, $\tau$ is the decay time constant, and $C$ is the steady-state component.

#### Statistical analysis of patch-clamp data

Data are presented as boxplots (median and boxes represent interquartile range) and mean ± standard error of the mean (SEM). Statistical analyses were conducted using SigmaStat 3.5 (Systat Inc, St. Louis, MO). Normality and equal variance assumptions were tested with the Kolmogorov-Smirnov and Levene median tests, respectively. For variant group comparisons with WT $K_v$4.3 currents, one-way ANOVA followed by the Holm–Šidák post hoc test was used for normally distributed data, while the Kruskal–Wallis test followed by Dunn's post hoc test was applied for non-normally distributed data. A p-value $< 0.05$ was considered statistically significant.

### Supplemental data

#### **Pathogenic *SCN5A* variants detected in *KCND3* carriers**

*Dutch BrS case with the KCND3:c.1798G>A/p.Gly600Arg variant*

The *SCN5A* p.Arg1628Gln variant is rare in the population (gnomADv4.1 filtering allele frequency = 0.0000039, PM2_Supporting), occurs in the fourth transmembrane region of *SCN5A* (PM1), is detected in multiple BrS cases (PS4_Supporting)[10] , is predicted to be deleterious (AlphaMissense score = 0.8128, PP3_Supporting) and was demonstrated to have severe loss-of-function in a calibrated high-throughput automated patch-clamp assay (z-score of -4.81, PS3) [11] . Classification = Likely Pathogenic (9 ACMG points).

*French BrS case with the KCND3:c.1859C>A/p.Pro620His variant*

The *SCN5A*:p.Arg893His is rare in the population (gnomADv4.1 filtering allele frequency = 0.00000072, PM2_Supporting), occurs in the second pore loop of *SCN5A* (PM1), is detected in multiple BrS cases (PS4_Moderate)[10] , is predicted to be deleterious (AlphaMissense score = 0.9758, PP3_Moderate) and was demonstrated to have severe loss-of-function in the automated patch-clamp assay (z-score of -5.4, PS3) [11] . Classification = Pathogenic (11 ACMG points).

#### Supplemental figures and tables

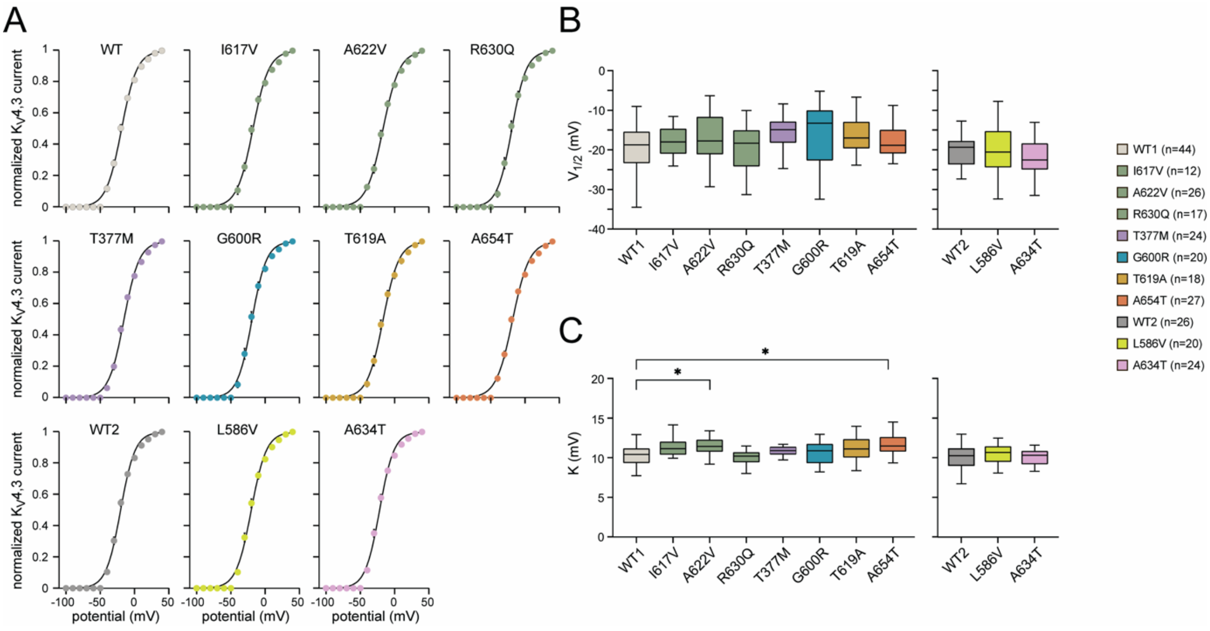

**Supplemental Figure 1. Effects of *KCND3* variants on the voltage dependence of $K_V$4.3 activation. (A)** Average voltage dependence of steady-state activation of WT and variant $K_V$4.3 channels. Solid lines represent Boltzmann fits. **(B)** Box plots of $V_{1/2}$ for activation. **(C)** Box plots of $K$ for activation, with an increased $K$ in A622V and A654T. *p < 0.05, Kruskal-Wallis test followed by Dunn's post hoc test.

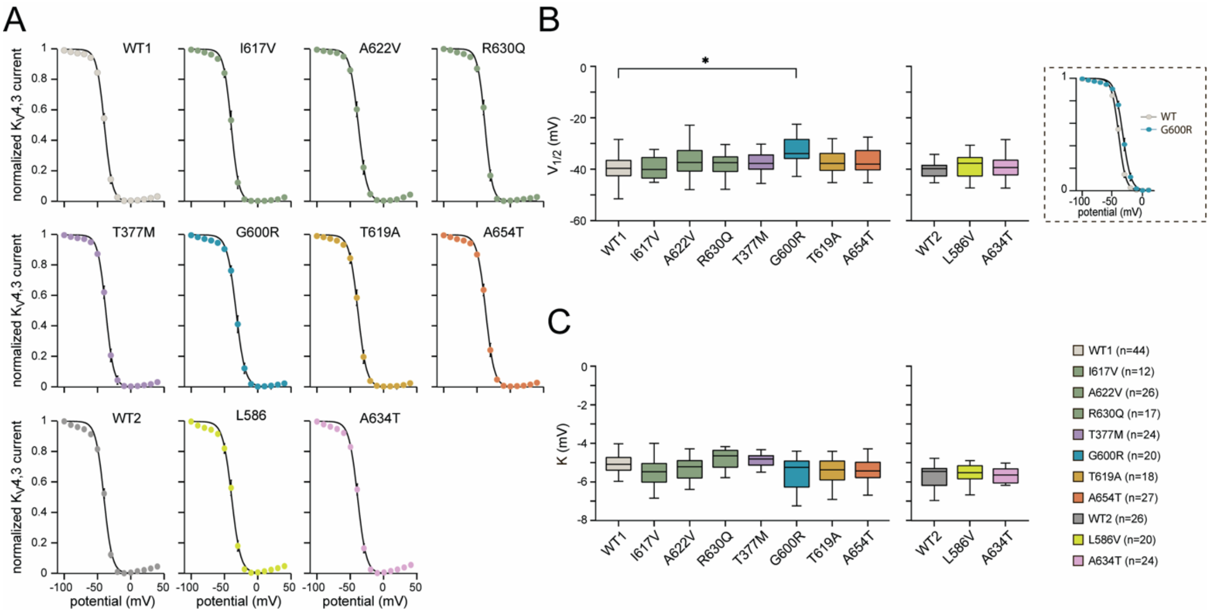

**Supplemental Figure 2. Effects of *KCND3* variants on the voltage dependence of $K_v$4.3 inactivation**. **(A)** Average voltage dependence of steady-state inactivation of WT and variant $K_v$4.3 channels. Solid lines represent Boltzmann fits. **(B)** Box plots of $V_{1/2}$ for inactivation. Inset shows representative steady-state inactivation curves from WT and p.G600R with a rightward shift of the inactivation curve in p.G600R. **(C)** Box plots of $K$ for inactivation.*$p < 0.05$, Kruskal-Wallis test followed by Dunn's post hoc test.

A

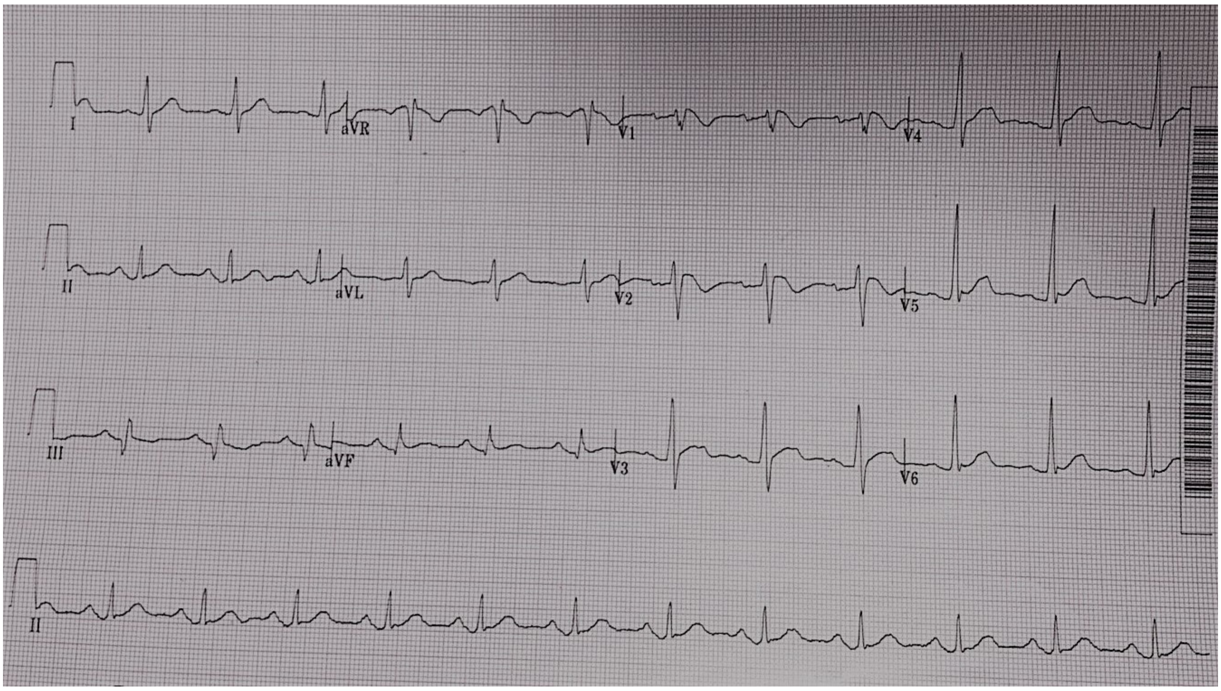
I
aVR
V1
V4
II
aVL
V2
V5
III
aVF
V3
V6
II

B

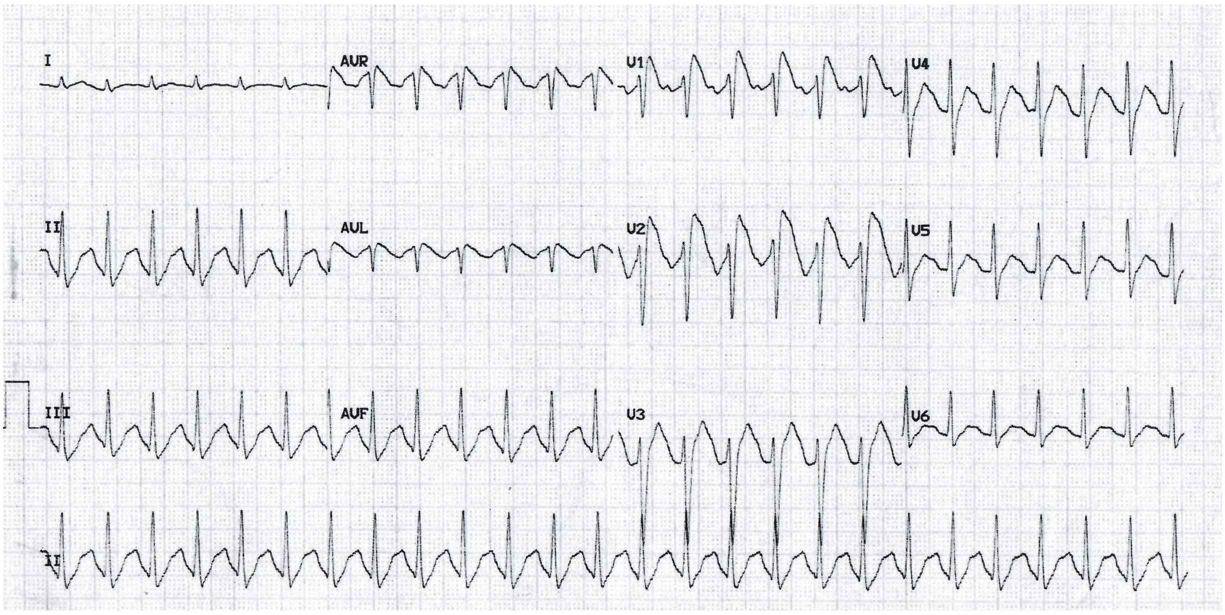
I
AVR
V1
V4
II
AVL
V2
V5
III
AVF
V3
V6
II

**C**

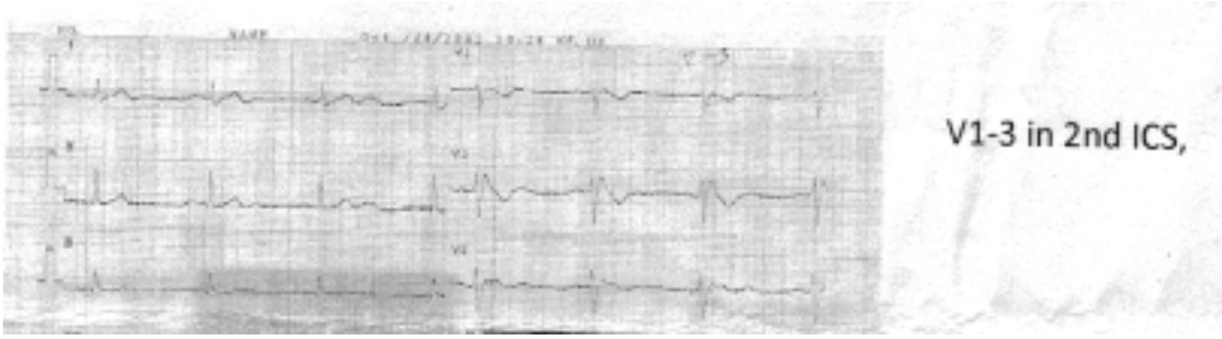

**D**

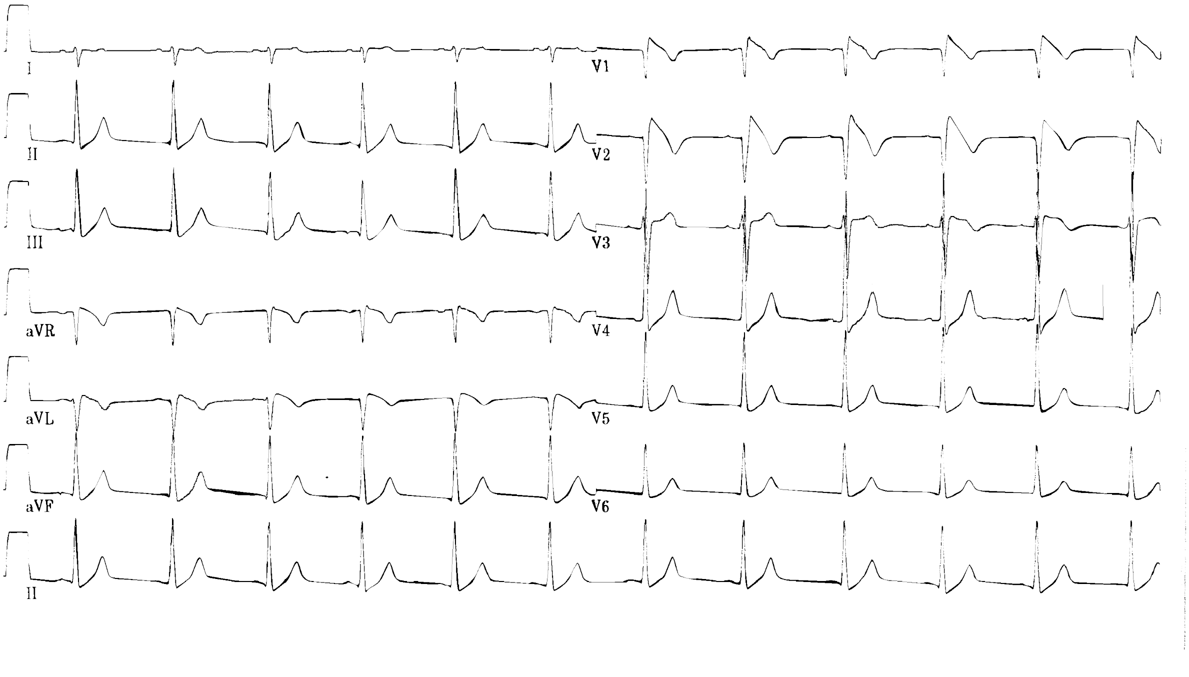

**E**

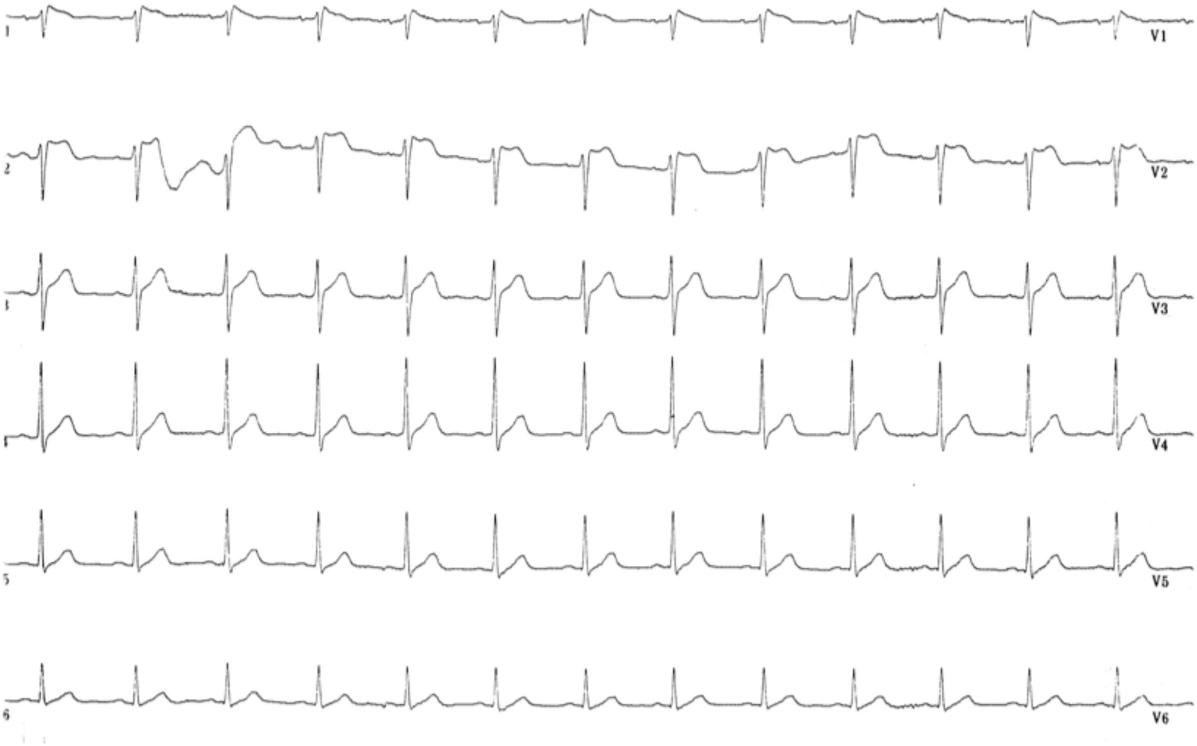

**F**

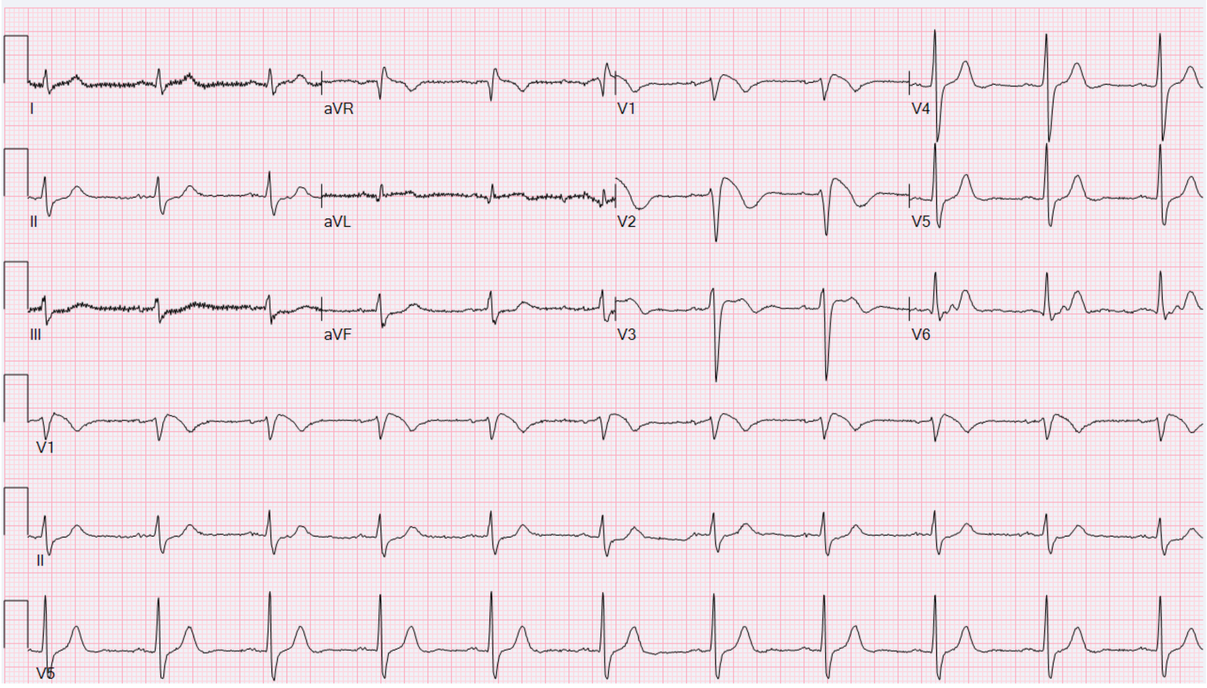

**Supplemental Figure 3. ECGs of BrS patients carrying functionally validated gain-of-function variants in *KCND3*.** **(A)** Type 1 BrS ECG pattern induced during sodium channel blocker challenge in Thai Patient 1 with the *KCND3*-p.Ala654Thr variant. **(B)** Spontaneous Type 1 BrS ECG pattern in Thai Patient 2 with the *KCND3*-p.Ala654Thr variant. **(C)** Spontaneous Type 1 BrS ECG pattern in Thai Patient 3 with the *KCND3*-p.Ala654Thr variant. **(D)** Spontaneous Type 1 BrS ECG pattern in the French patient carrying the *KCND3*-p.Ala634Thr variant. **(E)** Drug-induced BrS pattern in the French patient with the *KCND3*-p.Leu586Val variant. **(F)** Spontaneous Type 1 BrS ECG pattern in Dutch patient with the *KCND3*-p.Gly600Arg variant.

**Table S1.** List of rare (gnomAD v4.1 filtering allele frequency, FAF, <0.0001) *KCND3* missense variants detected in BrS case and control cohorts in this study. Variants are described with respect to the ENST00000302127 / NM_001378969 transcript, along with the location in the seven coding exons. Cohorts: European cases (n=752), European controls (n=1827), Thai cases (n=231), Thai controls (n=500).

| **Variant-HGVSc** | **Variant-HGVSp** | **Exon** | **Cases** | **Controls** | **Cohort** | **gnomAD v4.1_FAF** |
|---|---|---|---|---|---|---|
| c.145C>T | p.Arg49Trp | 1 | 0 | 2 | Thai | 0 |
| c.520G>A | p.Glu174Lys | 1 | 0 | 1 | European | 2.80E-07 |
| c.835_837del | p.Asn279del | 1 | 0 | 1 | European | 0 |
| c.1330A>G | p.Ser444Gly | 3 | 0 | 1 | Thai | 0 |
| c.1369A>G | p.Thr457Ala | 3 | 0 | 2 | Thai | 5.79E-05 |
| c.1502G>A | p.Arg501Gln | 5 | 0 | 1 | Thai | 1.57E-05 |
| c.1562T>C | p.Met521Thr | 6 | 0 | 1 | European | 0 |
| c.1646G>A | p.Arg549His | 6 | 0 | 1 | European | 0 |
| c.1756C>G | p.Leu586Val | 6 | 1 | 0 | European | 0 |
| c.1798G>A | p.Gly600Arg | 7 | 1 | 1 | European | 9.69E-05 |
| c.1855A>G | p.Thr619Ala | 7 | 1 | 0 | Thai | 0 |
| c.1859C>A | p.Pro620His | 7 | 1 | 0 | European | 8.03E-06 |
| c.1889G>A | p.Arg630Gln | 7 | 0 | 1 | Thai | 4.28E-05 |
| c.1900G>A | p.Ala634Thr | 7 | 1 | 0 | European | 0 |
| c.1960G>A | p.Ala654Thr | 7 | 3 | 0 | Thai | 1.77E-05 |

**Table S2.** Enrichment of rare (gnomAD v4.1 filtering allele frequency < 0.0001) *KCND3* missense variants in BrS case cohorts versus study controls and gnomAD population datasets, for the full transcript (ENST00000302127 / NM_001378969) and the final exon only. Analysis for complete cohort (European and Thai) and separately by ancestry using relevant gnomAD subpopulations (NFE - non-Finnish European, EAS - East Asian).

| Cohorts | Total samples | All of *KCND3* | | | Exon 7 only | | |
|---|---|---|---|---|---|---|---|
| | | Variants | Frequency | p value | Variants | Frequency | p value |
| All BrS | 983 | 8 | 0.81% | - | 7 | 0.71% | - |
| All Controls | 2327 | 12 | 0.52% | 0.33 | 2 | 0.09% | 0.004 |
| gnomAD_ALL | 728290 | 2321 | 0.32% | 0.015 | 600 | 0.08% | 2.0E-06 |
| European BrS | 752 | 4 | 0.53% | - | 3 | 0.40% | - |
| European Controls | 1827 | 5 | 0.27% | 0.296 | 1 | 0.05% | 0.077 |
| gnomAD_NFE | 554641 | 1731 | 0.31% | 0.3 | 395 | 0.07% | 0.017 |
| Thai BrS | 231 | 4 | 1.73% | - | 4 | 1.73% | - |
| Thai Controls | 500 | 7 | 1.40% | 0.75 | 1 | 0.20% | 0.037 |
| gnomAD_EAS | 19771 | 65 | 0.33% | 0.008 | 22 | 0.11% | 0.0002 |

**Table S3.** Demographic, genetic and clinical details of the family pedigree of the BrS patient carrying the *SCN5A*:p.Arg1628Gln and *KCND3*:p.Gly600Arg variants.

| **Pedigree ID** | **Sex** | **Age range (current)** | **Relationship to proband** | ***SCN5A* variant** | ***KCND3* variant** | **rs12090194** | **Clinical phenotype** |
|---|---|---|---|---|---|---|---|
| II-4 | M | 61-65 years | Proband | p.Arg1628Gln | p.Gly600Arg | CT | Baseline ECG showed a type 2 Brugada pattern; spontaneous type 1 Brugada ECG pattern was observed repeatedly during follow-up. Exercise testing revealed a wide-complex tachycardia with RBBB morphology and left axis deviation, followed during recovery by sinus rhythm with 5-mm coved ST-segment elevation. Polymorphic VT was inducible during electrophysiological study. An ICD was implanted. Normal cardiac MRI. |
| II-3 | M | 61-65 years | Brother | p.Arg1628Gln | p.Gly600Arg | CT | Asymptomatic but positive ajmaline test (monomorphic ventricular ectopy with LBBB morphology). Normal MRI, exercise test and Holter. |
| II-1 | F | 66-70 years | Sister | p.Arg1628Gln | p.Gly600Arg | CT | Negative ajmaline test and normal exercise test. Has vasovagal syncope. |
| II-2 | F | 66-70 years | Sister | WT | p.Gly600Arg | TT | Positive ajmaline test, vasovagal syncope. Normal Holter and exercise test. |
| I-2 | F | 96-100 years | Mother | WT | WT | CT | Unknown. |
| III-1 | F | 21-25 years | Daughter | WT | p.Gly600Arg | CT | Normal exercise test and echo but ECG revealed iRBBB with no ST elevations. Ajmaline test not performed. |

**Table S4.** Allele and genotype counts for the rs12090194 SNP in the UK Biobank and *All of Us* datasets, for non-Finnish European (NFE) individuals only.

Data shown for all individuals and separately for carriers of the *KCND3*:p.Gly600Arg variant.

| | **Alleles** | | | **Genotypes** | | | |
|---|---|---|---|---|---|---|---|
| **UK Biobank** | **AN** | **AC** | **AF** | | **Hom ref** | **Het** | **Hom alt** |
| All NFE | 917622 | 294003 | 0.320 | Counts | 211592 | 200435 | 46784 |
| | | | | Proportion | 0.461 | 0.437 | 0.102 |
| p.Gly600Arg NFE | 190 | 100 | 0.526 | Counts | 18 | 54 | 23 |
| | | | | Proportion | 0.189 | 0.568 | 0.242 |
| ***All of Us*** | **AN** | **AC** | **AF** | | **Hom ref** | **Het** | **Hom alt** |
| All NFE | 446690 | 138876 | 0.311 | Counts | 106124 | 95566 | 21655 |
| | | | | Proportion | 0.475 | 0.428 | 0.097 |
| p.Gly600Arg NFE | 76 | 47 | 0.618 | Counts | 1 | 27 | 10 |
| | | | | Proportion | 0.026 | 0.711 | 0.263 |
